# Impact of interventions on mpox transmission during the 2022 outbreak in Canada: a mathematical modeling study of three different cities

**DOI:** 10.1101/2024.06.20.24309262

**Authors:** Fanyu Xiu, Carla Doyle, Jorge Luis Flores Anato, Jesse Knight, Linwei Wang, Joseph Cox, Daniel Grace, Trevor A. Hart, Terri Zhang, Shayna Skakoon-Sparling, Milada Dvorakova, Rita Shahin, Herveen Sachdeva, Nathan Lachowsky, Hind Sbihi, Darrell H.S. Tan, Michael A. Irvine, Sharmistha Mishra, Mathieu Maheu-Giroux

## Abstract

**Background:** The 2022-2023 global mpox outbreak primarily affected gay, bisexual, and other men who have sex with men (GBM). It was met with swift community and public health responses. The relative impact of GBM’s reductions in sexual partners, contact tracing/isolation, and vaccination on transmission in Canadian cities remain unknown.

**Methods:** We estimated changes in sexual behaviours during the outbreak using 2022 data from the *Engage Cohort Study* which recruited self-identified GBM in Montréal, Toronto, and Vancouver (n=1,445). The numbers of sexual partners in the past 6 months (P6M) were modeled using negative binomial regressions. A transmission-dynamic compartmental model was calibrated to surveillance data. We estimated the averted fraction of new infections attributable to reductions in sexual partners, contact tracing/isolation, and first-dose vaccination, versus an unmitigated epidemic scenario, in each of the three cities.

**Results:** The empirical estimates of sexual behaviours changes were imprecise: 20% (RR=0.80; 95% credible intervals [95%CrI]: 0.47-1.36) fewer sexual partners among those reporting ≤7 partners (P6M) and 33% (RR=0.67; 95%CrI: 0.31-1.43) fewer among those with >7 partners (P6M). Compared to the unmitigated epidemics, we estimated that the three interventions combined avert 46%-58% of cases. Reductions in sexual partners, contact tracing/isolation prevented approximately 12% and 14% of cases, respectively. Vaccination’s effect varied across cities by start date and coverage, with 21%-39% mpox infections prevented.

**Conclusions:** Reduction in sexual activity, contact tracing/isolation, and vaccination all contributed to accelerating epidemic control and infections averted. Early vaccination had the largest impact.

## Background

The 2022-2023 global mpox outbreak resulted in more than 90,000 infections across 110 historically non-endemic regions, including Canada; it also disproportionately impacted gay, bisexual, and other men who have sex with men (GBM) [1]. Unlike historically reported cases in Central and West Africa, the recent global outbreak was sustained due to human-to-human transmission, primarily via sexual contacts [2]. From May 2022 to October 2023, 98% of 1,443 confirmed cases with available data in Canada, occurred among GBM [3]. Over 70% of the reported cases were concentrated in Montréal (Québec), Toronto (Ontario), and Vancouver (British Columbia), the country’s three largest cities [4–6]. Mpox’s cumulative incidences among GBM were around 1% in all three cities, despite differences in timing of outbreaks and interventions [7].

On May 19^th^, 2022, the first confirmed mpox case in Canada was reported in Montréal, the site of the first North American outbreak [8]. Cases in Toronto and Vancouver were respectively confirmed first on May 26^th^ and June 6^th^ of that year [8,9]. The outbreaks were met with swift responses from community organizations, sexual health professionals, laboratorians, and public health teams. The number of mpox cases peaked in all three cities from late-June to mid-July 2022 and declined thereafter, alongside the roll-out of public health interventions [3]. Various factors that govern the underlying transmission dynamics could have contributed to reductions in transmission, including saturation of groups at high risk of infection [10–13], changes in sexual behaviours following community outreach [14–16], contact tracing and isolation of traced contacts by local public health units [17,18], and use of vaccination [19,20].

Previous work showed that the effective reproduction number (i.e., *_t_:* the average number of secondary infections from one infectious individual where some people are no longer susceptible) could drop below 1 if even a small proportion (<2%) of GBM with the highest levels of sexual activity acquired immunity [7,11]. In Canada, public health authorities partnered with community-based organizations to amplify messaging about mpox prevention by reducing sexual partner numbers, prevention, testing, and vaccination on digital platforms and at gathering places [21,22]. Online surveys among convenience samples of GBM in the United Kingdom and the United States found that nearly half of interviewed GBM reported reducing their sexual partner numbers and visits to sex-on-premises venues after the onset of the mpox outbreak [15,16]. It remains uncertain if, and to which extent, GBM living in Canada similarly adapted their sexual behaviours in response to the mpox outbreaks, and what impact this had on the course of the epidemics.

Local health authorities conducted case and contact management to identify source infections and encouraged exposed symptomatic contacts to self-isolate [23]. In Montréal, 20% of contacts of confirmed cases were successfully traced/notified since late May 2022, resulting from the high number of anonymous sexual contacts [4,24]. Identified contacts were advised to self-monitor for symptoms and based on exposure risks, advised to receive post-exposure prophylaxis (PEP) using the Modified Vaccinia Ankara-Bavarian Nordic (MVA-BN) vaccine [4,23,24].

MVA-BN is a third-generation smallpox vaccine that offers cross-protection against the mpox virus [25,26]. In early June 2022, one-dose of vaccine for pre-exposure prophylaxis (PrEP) became available to individuals at high risk of exposure [5,24,27], including GBM who had sex with more than one partner and engaged in sexual contact in sex-on-premises venues [28]. As of mid-October 2022, approximately 24,000 first-doses in Montréal, and 35,000 in Toronto, of MVA-BN vaccines were administered [24,27]. In Vancouver, 18,000 first- and second-doses had been given over that same period [5].

Since mid-November 2022, case activity has been sporadic [3]. Understanding the impact of the main interventions in Montréal, Toronto, and Vancouver could generate new evidence, inform mpox prevention, and prioritize future public health actions. We aimed to evaluate the relative contribution of a) changes in sexual partner numbers, b) contact tracing/isolation, and c) first-dose vaccination on 2022 mpox outbreak dynamics among GBM in three different cities, while considering potential saturation of infections within high-risk groups. Specifically, we leveraged the *Engage Cohort Study,* a population-based study of GBM in the three cities, to empirically evaluate changes in sexual partner numbers during the period of high mpox transmission. We then developed a risk-stratified dynamic model of mpox transmission calibrated to mpox case surveillance data, to retrospectively assess the impact of various interventions on the final epidemic size, disentangling their unique contributions.

## Methods

### Data Source

The *Engage Cohort Study* is a prospective cohort of GBM in Montréal, Toronto, and Vancouver. Detailed descriptions of *Engage* can be found elsewhere [29–32]. Briefly, eligible participants were self-identified cisgender or transgender men living in one of the three cities, aged ≥16 years, reported sex with another man in the past 6 months (P6M), understood English and/or French, and provided written informed consent [33]. Participants were recruited during 2017-2019 in each city using respondent-driven sampling (RDS) with subsequent study follow-up visits every 6-12 months [34]. At each visit, participants completed an online questionnaire including questions about sexual behaviours.

### Changes in number of sexual partners during the mpox epidemics

To estimate changes in self-reported number of all-type sexual partners, we first excluded visits prior to 2022, given disruptions in sexual behaviours during the COVID-19 pandemic in 2020 and 2021 [7,35,36]. We defined the period over which mpox-driven behaviour changes could have occurred from May 19^th^, 2022 (first reported mpox case in Canada) to August 14^th^, 2022 (vaccination coverage >30% in all three cities). The latter date was chosen to ensure a reasonable sample size and account for mpox vaccination scale-up. Using data from all participants who had at least one visit in 2022, we fit a Bayesian negative binomial regression model to the number of sexual partners in the P6M, with a random intercept for each participant. We defined our exposure as the continuous fraction of the 6-month recall period that overlapped with the period of potential behaviour changes (i.e., adjusting for attenuation given the long recall period). We also included the following covariates: age (16-29, 30-39, 40-49, 50-59, ≥60 years), relationship status history (single/exclusive/open/unclear relationship at latest visit before 2022), HIV status (positive, negative/unknown), calendar month (continuous), and sexual partnership history (≤7 or >7 male sexual partners in the P6M at the latest visit before 2022). We then computed the relative change in sexual partner numbers (rate ratio, RR) during the mpox outbreak by sexual partnership history, to examine potential effect modification. Details on variable definitions and the regression model are available in the ***Supplementary Methods***.

To assess the sensitivity of our results, we repeated the analysis using alternative categorizations of sexual partnership history (≤3 or >3, and ≤5 or >5 male sexual partners in the P6M). We used a different end point of the behaviour change period (July 14^th^, 2022) to assess the sensitivity to an alternative exposure definition. We used a Bayesian logistic regression model to study two alternate outcomes: visit to bathhouses and/or sex clubs at least once in the P6M (binary) and attendance at group sex events at least once in the P6M (binary).

All empirical data analyses were performed with *R* (4.3.2) [37], using the *RStan* (2.32.3) [38] and *rstanarm* (2.26.1) [39] packages.

### Dynamic model of mpox transmission

We developed a dynamic, deterministic compartmental model of mpox transmission and control among a closed population of GBM. Surveillance data suggest that 30-39-year-olds, high numbers of sexual partners, and living with HIV were associated with mpox diagnosis [3,40]. We stratified the model into 5 age groups (16-29, 30-39, 40-49, 50-59, ≥60 years), 10 sexual activity groups (representing 60%, 90%, 95%, 96%, 97%, 98%, 99%, 99.5%, and 99.8% percentiles in the distribution of P6M sexual partner numbers), and HIV status (positive, negative/unknown). All GBM are assumed susceptible at the start of the outbreak, and they can acquire mpox and transition into the exposed (but not yet infectious) compartment, depending on a time-varying force of infection (***Figure 1***). The latter considers the mixing between GBM by age, sexual activity, and HIV status. The age and HIV mixing were informed by previous studies [41], and degree of assortativity by sexual activity was calibrated. Based on a previous analysis [7], we allowed for underreporting and reporting delays [42,43]: 77%-86% of all infections were estimated to be reported in the surveillance data.

**Figure 1.**
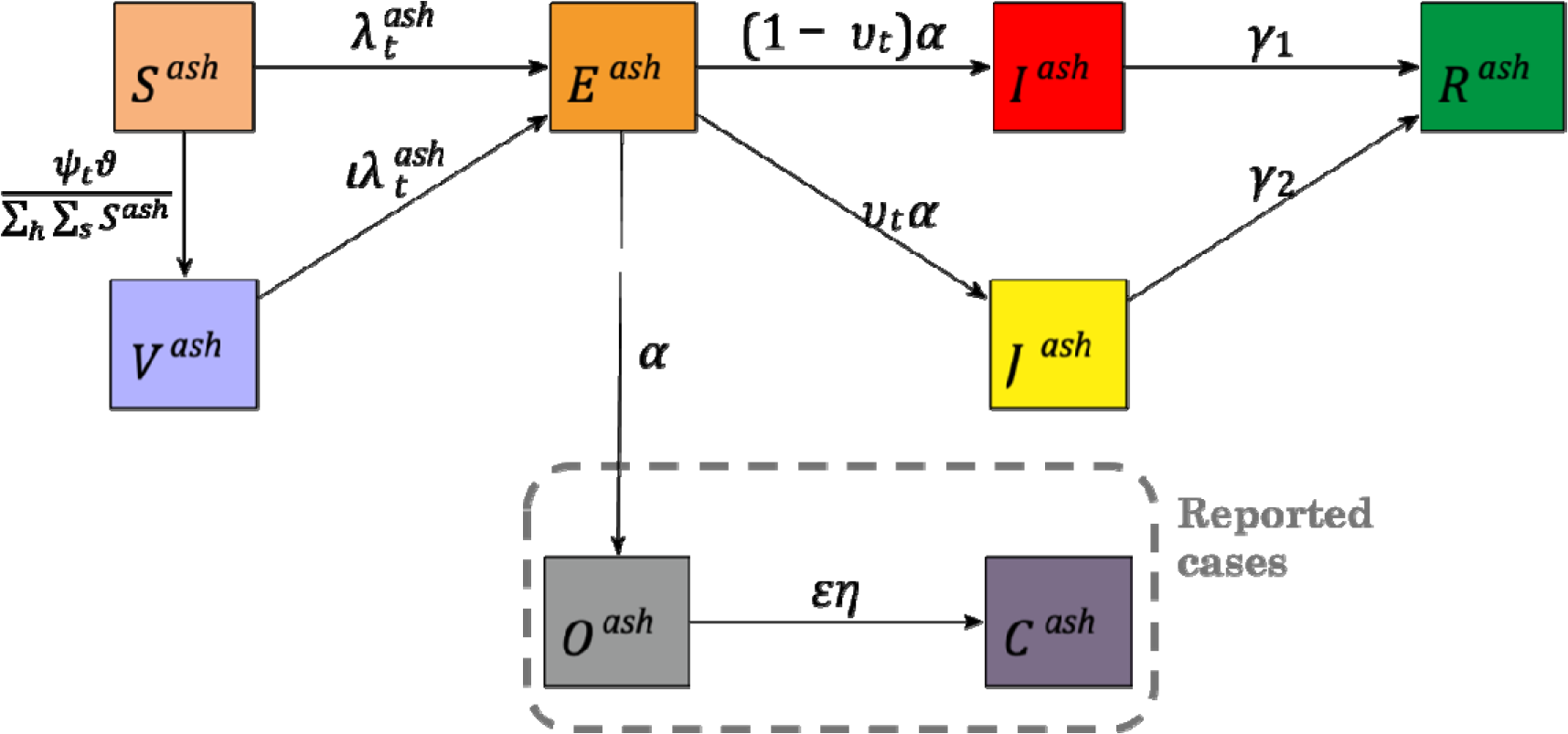
Diagram of the compartmental flows of the deterministic model of mpox virus transmission among gay, bisexual, and other men who have sex with men (GBM). *a, s, h*: superscripts for age groups, sexual activity groups, and HIV status, respectively. The name of the compartments refers to susceptible (S), exposed (E), infectious (I), removed (R), vaccinated (V), and isolated (J). Removed refers to the state where GBM are no longer infectious, stopped having sex, or developed natural immunity to mpox. Two other compartments are used to track symptoms onset (O) and the case confirmation process (C). The main parameters are the following: ψ*_t_*: first-dose vaccination doses at time t; proportion of vaccinations received by age groups; 1-vaccine effectiveness (assuming leaky type); : force of infection specific to group *a, s, h* at time t; : rate of infectivity onset among exposed ≈ (latent period)^−1^; υ*_t_*: proportion traced and isolated among exposed at time t; : rate of removal among infectiou individuals who are not traced and isolated = (effective infectious period)^−1^; rate of removal among infectious individuals who are traced and isolated = (self-isolation period)^−1^; ∶ the reporting fraction (proportion of cases that are reported, calibrated parameter); ∶ reporting delay. Values of parameters are defined in *Table S2*.

In terms of interventions, community and public health messaging could have led to potential behaviour changes. GBM’s contact rates were allowed to vary using a rate ratio that reflected potential reductions in sexual activity (as informed by a prior from the empirical analysis above). Exposed GBM can be traced and isolated by local public health authorities. Local public health authorities in Montréal suggested that 20% of contacts of reported cases were traced [4,24]. We adjusted that 20% by the fraction of cases reported in each city and the fraction of contacts traced before being infectious. Given limited vaccine supply, first-dose vaccination coverage was maximized by delaying the administration of second doses [24]. We only modeled first-dose vaccination, as it constituted >90% of vaccines administered before mid-October 2022 in all three provinces [5,24,27]. We used the age-specific weekly doses administered from publicly available reports [4,5,24,27]. Vaccine effectiveness was modeled using a leaky-type vaccination compartment–partially effective for all vaccinated, instead of fully effective for a fraction vaccinated [44]. Values of model parameters are defined in ***Table S2***. Model structure is presented in ***Figure 1*** and details are in ***Supplementary Methods***.

### Model calibration

We calibrated the model using mpox surveillance data, assuming all reported cases were among GBM [3–6,24,45]. The model was calibrated jointly for the three cities using a Bayesian sampling importance resampling and a negative binomial likelihood for daily numbers of total reported cases.

We cross-validated our models to the following outcomes (whenever available): the proportion of mpox cases who were among people living with HIV in Montréal, the age distribution of cases in Montréal and Toronto, and the age distribution of vaccines received in Montréal and Toronto.

### Averted fraction of new infections from past interventions

Using the calibrated model, we evaluated the impacts on mpox transmission of 1) changes in number of sexual partners, 2) contact tracing/isolation, 3) first-dose vaccination, 4) all three measures combined, and 5) combinations of any two interventions. We estimated the impact using the cumulative averted fraction (AF) of new infections, calculated as the difference in the cumulative incidence between each of the interventions scenarios above divided by the counterfactual cumulative incidence corresponding to an unmitigated epidemic.

### Sensitivity analysis for the averted fraction of new infections

First, we assessed the model sensitivity to our prior for the changes in sexual partner numbers by fixing the RR to the point estimates found in the empirical analysis. Second, we reduced the proportion of contacts traced from 20% to 15% and 10% to reflect potential non-disclosure of sexual contacts. Third, we examined the sensitivity of our results to vaccine effectiveness by using the minimum and maximum of estimates from literature (35.8% and 86.0%, respectively) [46–52]. Finally, to directly compare the impact of vaccination in all three cities, we examined a scenario where vaccination was initiated the same number of days after detection of the first local cases and reached the same daily coverage, using Vancouver as the reference since it had an earlier start of vaccination and achieved the highest vaccine coverage. For the first three analyses, we re-calibrated the model and re-calculated the averted fractions.

The model was coded in R, using a C++ back-end [53], and solved using an Euler algorithm with a 6-hour time step. Additional details on methods can be found in the ***Supplementary Methods***.

## Results

### Changes in numbers of sexual partners during the mpox epidemics

Out of 1,957 visits from participants that occurred during 2022, 424 visits partly overlapped with the potential behaviour change period (May 19^th^-August 14^th^, 2022). Although imprecise, the results from the regression model were indicative of a decrease in sexual partners. Specifically, we estimated a RR of 0.80 (95% Credible interval [CrI]: 0.47-1.36) and 0.67 (95%CrI: 0.31-1.43) among those with >7 (P6M) and ≤7 sexual partners at their last visit before 2022 (***Table 1***). Summary statistics of the study population from *Engage* and the effect estimates of covariates are presented in ***Table S6*** and ***Table S7***. Sensitivity analyses are shown in ***Table S8***. The results on changes in visits to sex-on-premises venues and attendance of group sex were inconclusive.

**Table 1.** Changes in the number of reported sexual partners in the past 6 months during the period of mpox-driven potential behaviour changes among participants in the *Engage Cohort Study,* by sexual partnership history.

| Numbers of sexual partners at the latest visit before 2022 | Number of visits |  | RR (95%CrI) |
| --- | --- | --- | --- |
|  | *During Mpox outbreak* | *Rest of 2022* |  |
| $\leq 7$ sexual partners | 324 | 1211 | 0.80 (0.47, 1.36) |
| $> 7$ sexual partners | 100 | 322 | 0.67 (0.31, 1.43) |

There were 424 visits for which the past 6 months recall period overlapped with the mpox epidemic. Among those, an average of 19% of the recall period was within the epidemic period. Models adjusted for months since January 1^st^, 2022 (1, 2, …, 12; continuous), age (16-29, 30-39, 40-49, 50-59, ≥60 years), relationship status history (single, exclusive relationship, open relationship, unclear), and HIV status (binary). *CrI* = credible interval; *RR* = rate ratio.

### Model calibration

The city-specific models replicated the daily number of reported cases (***Figure 2***), with a slight overestimation of cases in Toronto during the Fall of 2022. Furthermore, the model-predicted age distribution and HIV status of cases generally matched well with the one reported from local surveillance data (***Figure S2***). The model slightly overestimated cases in the 16-29 age groups and underestimated those in the 30-39 groups in Montréal and Toronto. The posterior parameter distributions for calibrated parameters are in ***Table S9***.

**Figure 2.**
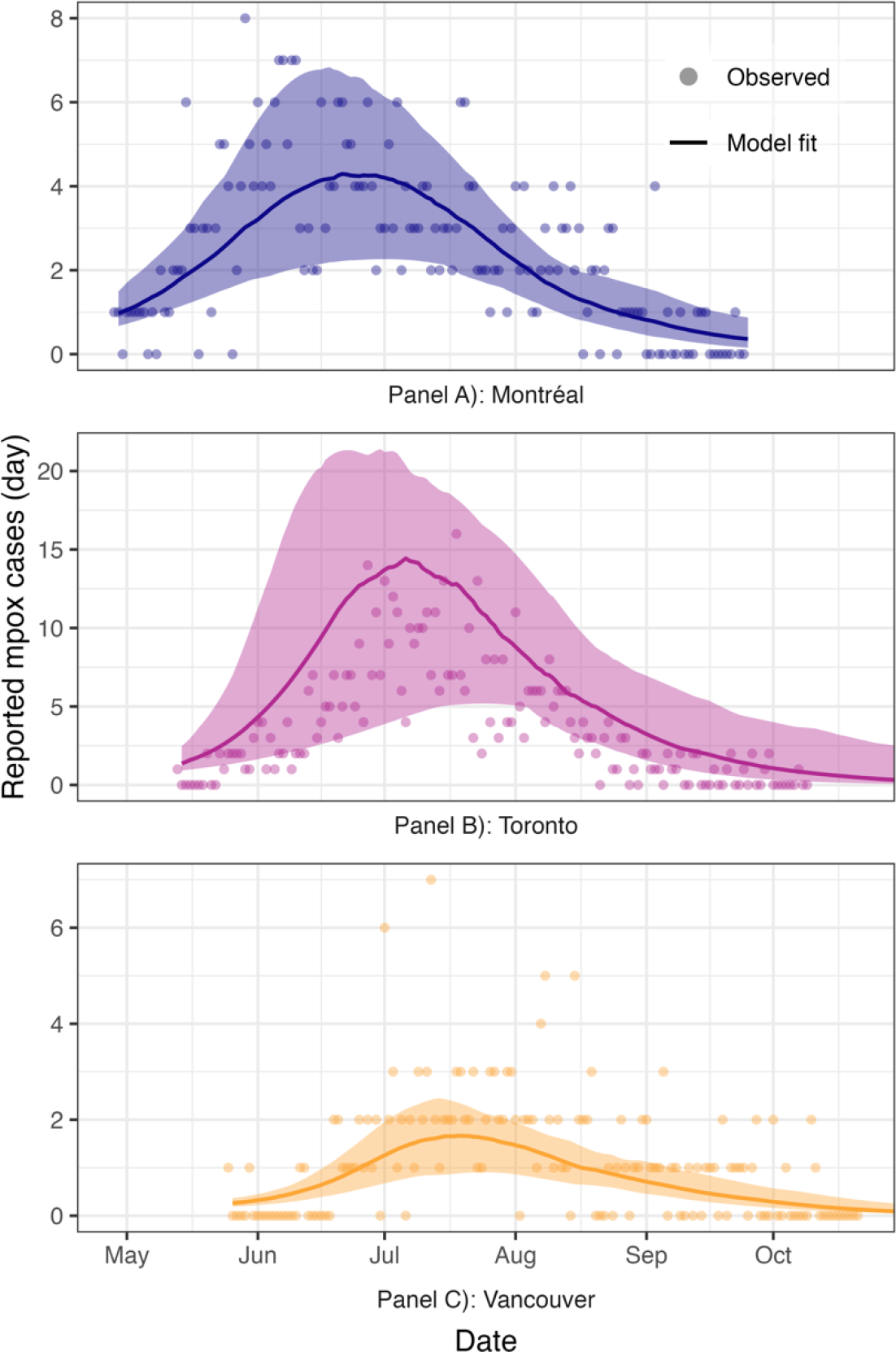
Model-calibrated epidemic curves and observed mpox case data in Montréal (A), Toronto (B), and Vancouver (C) during 2022. The reported case data (points) and model fits (curves) are presented up to 150 days after the first reported mpox cases in each city. The solid line is the median of the modeled cases, and the shaded area shows the 95% credible interval for the mean number of daily cases.

### Fraction of cases averted by interventions

In the three cities, we found that the outbreaks would have subsided without any interventions, but with nearly 50% more infections (***Figure 3***). Combined, we estimated that the three interventions averted 48% (35%-66%), 46% (34%-67%), and 58% (52%-69%) infections in Montréal, Toronto, and Vancouver, respectively. The calibrated RR for changes in number of sexual partners was 0.94 (95%CrI: 0.80-0.99) across sexual partnership history level defined by >7 partners and 0.94 (0.70-0.99) among those defined by ≤7 partners. The changes in number of sexual partners moderately decreased transmission: the estimated AF was 15% (3%-34%) in Montréal, 11% (2%-27%) in Toronto, and 10% (2%-22%) in Vancouver. Contact tracing averted 14% (12%-21%), 14% (12%-22%), and 14% (12%-16%) of infections in Montréal, Toronto, and Vancouver, respectively. We estimated that first-dose vaccine coverage among GBM reached 44% (Montréal), 45% (Toronto), and 58% (Vancouver) by mid-October 2022 (***Figure S3***). The impact of vaccination varied according to vaccine coverage and the timing of vaccine campaign initiation relative to the beginning of the outbreak. Vaccination averted 21% (16%-33%), 22% (16%-41%), and 39% (35%-48%) of infections in Montréal, Toronto, and Vancouver, respectively. The added effects of any combinations of two interventions were roughly equal to the sum of their individual effects (***Table S10)***.

**Figure 3.**
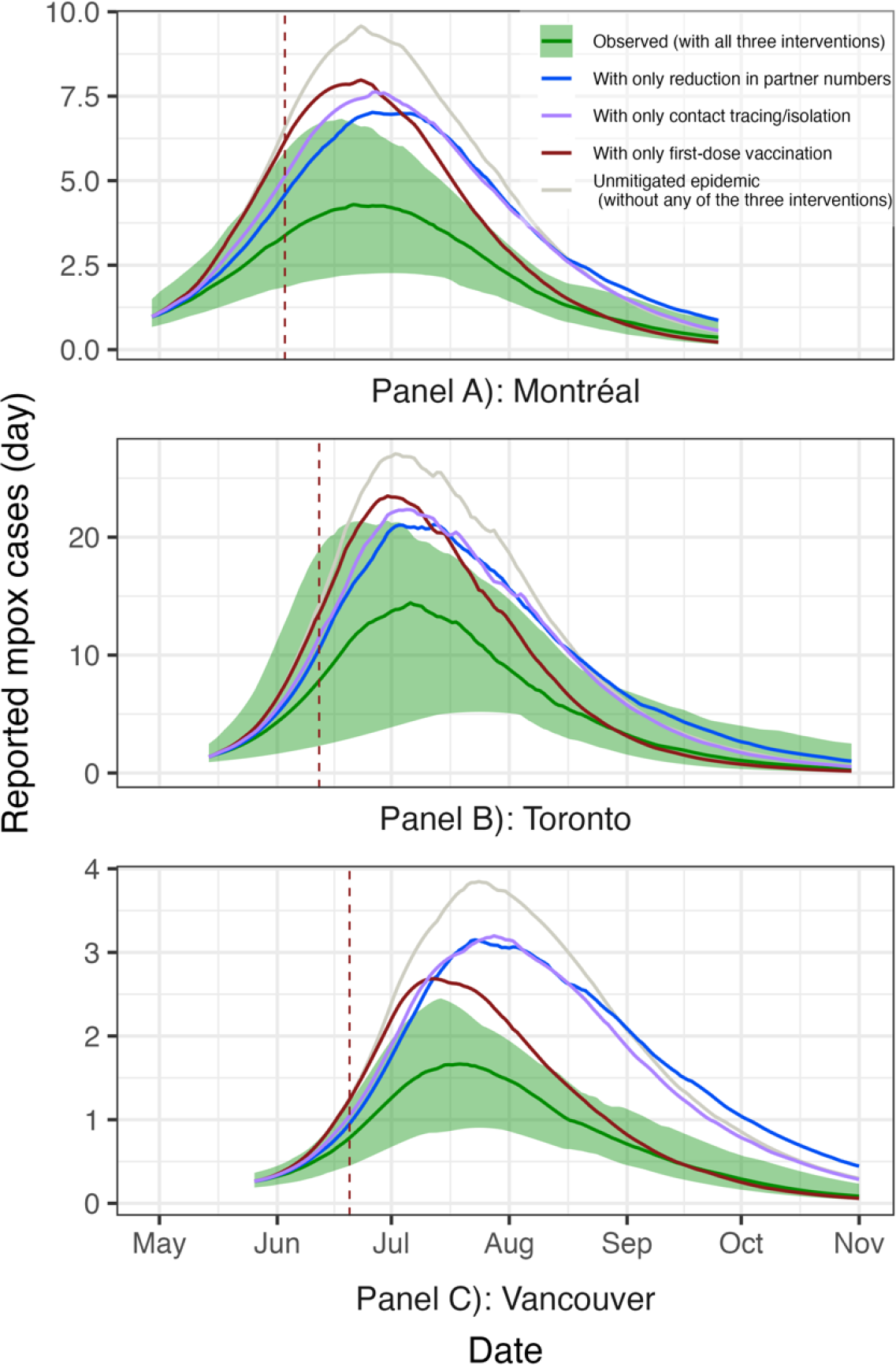
Model-predicted daily reported mpox cases in 2022 in Montréal, Toronto, and Vancouver under observed interventions levels and scenarios with selected interventions. The lines represent the median. The green lines and shaded area (95% credible intervals) correspond to the observed epidemic curves in the three cities. The grey lines are the modeled unmitigated epidemic. The blue lines are the epidemic curves with reduction in sexual partner numbers alone, the purple lines are with contact tracing/isolation alone, and the maroon lines are with first dose vaccination alone. Vertical dashed lines show the start of one-dose vaccination in each city: June 3^rd^, June 12^th^, and June 20^th^, 2022, in Montréal, Toronto, and Vancouver, respectively. The estimates for intervention scenarios were shown in *Table S10*.

### Sensitivity analyses for the fraction of cases averted by interventions

Fixing the values of the RR for the reduction in numbers of sexual partners to the point estimates from the empirical behavior change analysis did not replicate the epidemic well. This suggests that such parameters are not compatible with the observed outbreak trajectories. The fraction of cases averted by contact tracing was reduced to 10%-11% and 7% when using a proportion of 15% and 10% of cases traced, respectively (***Table S10***). First-dose vaccination prevented 14%-29% of cases when using 35.8% vaccine effectiveness (1-dose). Conversely, assuming 86.0% vaccine effectiveness, the number of cases prevented increased to 38%-59%. Finally, standardizing the start of vaccination and vaccine coverage to Vancouver resulted in a similar fraction of cases averted by vaccination in the three cities (38%-41%).

## Discussion

The 2022-2023 mpox outbreak was met with swift community and public health responses in Canada, inspired by GBM communities’ decades-long fight against HIV. Using cohort data from a representative sample of urban GBM in Canada and a calibrated risk-stratified dynamic model of mpox transmission, we estimated that, altogether, behaviour changes, contact tracing/isolation, and vaccination averted 46%-58% of cases among GBM in Canada’s three largest cities. Among these, vaccination had the largest impact on averted cases despite moderate coverage of first doses (44%-58%) among GBM. Vaccines alone averted an estimated 21%-39% of new infections, depending on the city. Our findings support implementing targeted vaccination quickly and at-scale if localized epidemics resurge and continuing immunization among GBM with multiple sexual partners. This measure is relevant given current low coverage of the second doses, higher vaccine effectiveness conferred by two-doses [47], current absence of vaccination clinics in the cities, and localized epidemic resurgence in Toronto in early 2024 [45].

Our analysis, based on a large, population-based cohort, suggests that GBM may have changed sexual behaviours to have fewer sexual partners during the mpox outbreak. However, the estimate was highly uncertain, particularly given the drop in sexual partner numbers that had already occurred due to previous COVID-19 lockdowns [36], precluding a definite conclusion from the empirical analysis. Nevertheless, our calibrated dynamic models suggest that small declines in partner numbers are compatible with the observed epidemics. These averted 10%-15% of cases across the three cities.

Assuming that 20% of contacts were traced (as reported from Montréal), tracing/isolation of exposed cases averted 14% of infections in the three cities. This impact was sensitive to the proportion of traced sexual contacts. Lack of contact information reported by cases, largely due to anonymous partnering, limited the proportion of contacts traced and isolated.

As of mid-October 2022, we estimated that the large-scale vaccination of first-dose of MVA-BN vaccine attained coverage of 44%, 45%, and 58% among GBM in Montréal, Toronto, and Vancouver, respectively [5,24,27]. Assuming 51.5% vaccine effectiveness, first-dose vaccination averted 39% of infection in Vancouver (95%CrI: 35%-48%), 22% in Toronto (95%CrI: 16%-41%) and 21% in Montréal (95%CrI: 16%-33%). High vaccine coverage in Vancouver may explain the higher impact in that city (***Table S10***).

Despite these notable impacts, the mpox outbreaks could have waned without any of the three interventions. This is consistent with the saturation of “core groups”, resulting in the accumulation of infection-derived immunity against mpox and, ultimately, the epidemic’s downturn. However, it is likely that, the community and public health responses greatly accelerated the decline in incidence, as has been found in other settings [10,12].

Our results should be interpreted considering several limitations. First, we had limited information on contact tracing/isolation and it is difficult to effectively model this intervention using compartmental models. We used an approximation to estimate the proportion of publicly traced cases that would be isolated before onset of infectiousness. However, case self-notification of partners was not captured by contact tracing data, which means we could have underestimated the impact of contact tracing/isolation. Second, we assumed that GBM had initially no prior immunity against mpox. However, some GBM born before smallpox vaccination stopped in Canada in 1972 and those that immigrated from certain countries could have previously received smallpox vaccines [54,55]. This should not change our results much because people aged >50 years old represented only <14% all cases in Canada [3]. Finally, vaccination was age-specific, but it did not fully reflect prioritization by partner numbers and HIV status, whereas individuals at higher risk of mpox acquisition may have been more likely to receive vaccines, meaning the effect of vaccination could have been underestimated.

The strengths of this study include the use of data from a large population-based cohort to inform model parameterization and statistical analyses. This approach allowed us to empirically explore the impact of behaviour changes. Second, we accounted for balanced mixing by age groups, sexual activity groups, and HIV status in the mpox transmission model, all strongly associated with mpox diagnoses in surveillance data [3,56]. Finally, we calibrated the model in a Bayesian framework, ensuring that parameter uncertainty is reflected in our model estimates, and cross-validated our model predictions.

## Conclusion

GBM in Montréal, Toronto, and Vancouver may have decreased sexual partnering during the transmission period of the 2022 mpox outbreak, which alongside contact tracing/isolation, contributed to averting mpox infections. Early vaccination was key to reducing the number of mpox infections. While mpox outbreaks in Canada could have eventually subsided without intervention, 50% more cases could have been infected, leading to unnecessary harms and potentially serious health consequences.

## Declarations

## Ethics

The Research Institute of the McGill University Health Centre and the Research Ethics Office of the Faculty of Medicine and Health Sciences, McGill University (A06-M32-23B), Toronto Metropolitan University (REB #2016-113), the University of Toronto (protocol #00033527), St. Michael’s Hospital (REB #17-043), the University of Windsor (REB #33443), the University of British Columbia (H16-01226), Providence Health Care (H16-01226), the University of Victoria (H16-01226), and Simon Fraser University (H16-01226) provided ethics approvals for the *Engage Cohort Study*. The Research Ethics Office of the Faculty of Medicine and Health Sciences at McGill University approved the secondary analyses of the data in this study.

## Data Availability

All data produced in the present work are contained in the manuscript.

https://github.com/pop-health-mod/mpox-intervention

## Acknowledgments

The Engage Cohort Study is led by Principal Investigators in Montreal by Joseph Cox and Gilles Lambert; in Toronto by Trevor A. Hart & Daniel Grace; and in Vancouver by Jody Jollimore, Nathan Lachowsky, and David Moore. More information about the Engage Cohort Study can be found here: https://www.engage-men.ca. The authors would like to thank the *Engage* study participants, office staff, and community engagement committee members, as well as our community partner agency, *RÉZO*.

## Author contribution

FX, CD, SM, MI, and MM-G contributed to the conception and design. JC, DG, TAH, MD, NJL, SSS, and SM were involved in the design, data collection, and data management of the *Engage Cohort Study*. Analyses were performed by FX, with support from CD, JLFA, and MM-G. LW, JK, MI, and SM provided input on preliminary methods discussions. JK and LW developed the preferential mixing matrices, and JK coded the algorithm. The manuscript was drafted by FX. All authors reviewed the manuscript for important intellectual content. Overall supervision for this project was provided by MM-G. All authors approved the final manuscript. The Engage study is led by the following principal investigators (in alphabetical order): Joseph Cox, Daniel Grace, Trevor Hart, Jody Jollimore, Nathan Lachowsky, Gilles Lambert, and David Moore.

## Software and source code

The model package and code for analyses are available on a GitHub repository (https://github.com/pop-health-mod/mpox-intervention).

