## Supplementary Materials for "Impact of interventions on mpox transmission during the 2022 outbreak in Canada: a mathematical modeling study of three different cities"

### Text S1: Supplementary information on methods and approaches used

#### *Changes in numbers of sexual partners during the mpox epidemics*

We fitted a Bayesian negative binomial regression model (with a random intercept for each participant estimated using partial pooling) to estimate the relative change (rate ratio, RR) in sexual partner numbers in the past 6 months (P6M) during the period of mpox-driven behaviour changes (May 19^th^, 2022-August 14^th^, 2022; 87 days). We chose a negative binomial likelihood since it best described GBM’s distribution of numbers of sexual partners in previous work [1]. Given the 6-month recall period for our outcome, we expect that the change in the log-number of sexual partners will be proportional to how much of the recall period overlaps with the mpox outbreak. To adjust for that effect attenuation, we defined the exposure variable as the fraction of the recall period that covers the period of mpox-driven behaviour changes, as follows:

$$\boldsymbol{x}\left( t \right)\boldsymbol{=}\left\{ \begin{aligned} 0 if t<t_{mpox_{start}} or t>t_{mpox_{end}} \\ \frac{\left( t-t_{mpox_{start}} \right)+1}{6 months} if t_{mpox_{start}}\leq t<t_{mpox_{end}} \end{aligned} \right.$$

where *t* is the visit date, $t_{mpox_{start}}$ is May 19^th^, 2022, and $t_{mpox_{end}}$is August 14^th^, 2022.

Given the short window of the defined period, we conducted a pooled analysis across the three cities to improve precision. Additionally, each participant $i$ has a unique random intercept to account for correlation between their multiple visits. Informed by the peer-reviewed literature [1–5], we included the following variables as covariates in the model: age group (16-29, 30-39, 40-49, 50-59, ≥60 years), relationship status history (single, exclusive relationship, open relationship, unclear), and sexual partnership history (≤7 or >7 sexual partners in the P6M at the latest visit before 2022), HIV status (binary), and calendar month since January 1^st^, 2022 (continuous) (***Table S1***). We included the last variable to account for potential secular trends in sexual activity among GBM in the aftermath of the COVID-19 pandemic [1]. A product term between the exposure and sexual partnership history (i.e., ≤7 or >7 male sexual partners in the P6M at the latest visit before 2022) was included to reflect potential heterogeneity in the effect of the mpox outbreak by sexual activity levels on the number of sexual partners.

The regression model was fitted using Hamiltonian Monte Carlo in *rstanarm*, with 4,000 iterations (2 chains, 2,000 burn-in iterations, no thinning, ensuring that the effective sample size for each parameter was ≥1,000 or larger). We used weakly informative priors and assessed convergence via traceplots and the potential scale reduction factor ($\hat{R}$). The regression models, with negative binomial likelihood (NB), can be written as

$$y_{it} \sim NB( \lambda_{it}, \phi)$$

$$\log(\lambda_{it})=\alpha_{0}+\mu_{i}+\beta X_{it}$$

where

$i:$ index for participants, $i\in\left\{ 1,2,\ldots,1445 \right\}$;

$y_{it}:$observed number of all-type sexual partners in the P6M for participant $i$ at time $t$;

$\lambda_{it}:$ mean model-predicted number of partners for participant $i$ at time $t$;

$\phi:$ overdispersion parameter;

$\alpha_{0}:$ fixed intercept;

$\mu_{i}:$ random intercept for participant $i$;

$X_{it}:$avector containing the values of predictors for participant $i$ at time $t$, i.e., *exposure variable*, *age, relationship status, HIV status, calendar month, sexual partnership history level, exposure* $\times$*sexual partnership history level.*

$\beta:$ vector of regression coefficients corresponding to the matrix of covariates $X$;

We used the following prior distributions for the regression parameters. Note the second argument refers to the standard deviation.

$$\alpha_{0}\sim Normal\left( 0, 10 \right)$$

$$\beta_{k}\sim Normal\left( 0, 10 \right)$$

$\forall k\in${*exposure variable*, *age, relationship status, HIV status, calendar month, sexual partnership history level, exposure* $\times$*sexual partnership history level*}

$$\phi\sim halfCauchy\left( 0, 5 \right)$$

The covariance matrix of the random intercepts can be decomposed into the correlation matrix and variances:

$$\left[ \begin{matrix} \sigma_{\mu_{1}}^{2} & \cdots& \rho_{\mu_{1},\mu_{1445}}\sigma_{\mu_{1}}\sigma_{\mu_{1445}} \\ \vdots& \ddots& \vdots\\ \rho_{\mu_{1},\mu_{1445}}\sigma_{\mu_{1}}\sigma_{\mu_{1445}} & \cdots& \sigma_{\mu_{1445}}^{2} \end{matrix} \right]=\left[ \begin{matrix} \sigma_{\mu_{1}} & \cdots& 0 \\ \vdots& \ddots& \vdots\\ 0 & \cdots& \sigma_{\mu_{1445}} \end{matrix} \right]\left[ \begin{matrix} 1 & \cdots& \rho_{\mu_{1},\mu_{1445}} \\ \vdots& \ddots& \vdots\\ \rho_{\mu_{1},\mu_{1445}} & \cdots& 1 \end{matrix} \right]\left[ \begin{matrix} \sigma_{\mu_{1}} & \cdots& 0 \\ \vdots& \ddots& \vdots\\ 0 & \cdots& \sigma_{\mu_{1445}} \end{matrix} \right]$$

The priors for the correlation matrix and variances are:

$$\left[ \begin{matrix} 1 & \cdots& \rho_{\mu_{1},\mu_{1445}} \\ \vdots& \ddots& \vdots\\ \rho_{\mu_{1},\mu_{1445}} & \cdots& 1 \end{matrix} \right]\sim LKJcorr(1)$$

$$\sigma_{\mu_{i}}\sim exponential\left( 1 \right) \forall i\in\{1,2,\ldots,1445\}$$

The $LKJcorr(1)$ is the Lewandowski-Kurowicka-Joe distribution with shape parameter equals to 1.

#### *Structure for the dynamics model of mpox transmission and control*

We modelled transmission in Montréal, Toronto, and Vancouver, using separate but jointly calibrated compartmental models, for 150 days after first confirmed cases in each respective city [6,7]. The modelled period covered the majority of cases in the outbreaks, after which only sporadic cases were observed in the three cities [8–10]. The model considers the degree of assortativity in sexual mixing between GBM by age, sexual activity, and by HIV status. Given the short timeframe of the mpox outbreaks, we did not model HIV dynamically and assumed a closed population (that is, individuals did not enter or exit the modelled population during the outbreak period). The model considers that all GBM were equally susceptible to mpox at the beginning of the outbreak. Given reports of asymptomatic cases and case underreporting [11], we assumed that only a 77%-86% of all infections was reported in the surveillance data based a previous modelling study [1]. Finally, we accounted for an average 2-day delay between symptom onset and case confirmation reported in the surveillance data [12].

Susceptible GBM can acquire mpox and transition into the exposed (but not yet infectious) compartment, depending on a time-varying force of infection (***Figure 1***). After an average of 5.1 days [13–17] (***Table S2***), exposed GBM become infectious but a time-varying proportion can isolate if traced by local public health authorities (due to limited data, we assumed isolation only came from case contacts traced by public health agencies). In the infectious stage, people will remain infectious until they recover (no longer infectious) or stop sexual activity due to mpox symptoms (i.e., the effective infectious period, to be calibrated).

#### *Equations for the dynamics model of mpox transmission and control*

The system of ordinary differential equations describing mpox’s transmission dynamics is presented below. GBM in each city are partitioned into age groups *a*, sexual activity level *s*, and HIV status *h*. The natural history parameters are presented in ***Table S2***.

$$\frac{dS^{ash}\left( t \right)}{dt}=-\lambda_{t}^{ash}S^{ash}\left( t \right){-\psi}_{t}\vartheta^{a}\frac{S^{ash}\left( t \right)}{\sum_{h} \sum_{s} S^{ash}(t)}$$

$$\frac{dV^{ash}\left( t \right)}{dt}=\psi_{t}\vartheta^{a}\frac{S^{ash}\left( t \right)}{\sum_{h} \sum_{s} S^{ash}(t)}-\iota\lambda_{t}^{ash}V^{ash}\left( t \right)$$

$$\frac{dE^{ash}\left( t \right)}{dt}=\lambda_{t}^{ash}S^{ash}\left( t \right)+\iota\lambda_{t}^{ash}V^{ash}\left( t \right)-\alpha E^{ash}\left( t \right)$$

$$\frac{dI^{ash}\left( t \right)}{dt}=\left( 1-\upsilon_{t} \right)\alpha E^{ash}\left( t \right)-\gamma_{1}I^{ash}\left( t \right)$$

$$\frac{dJ^{ash}\left( t \right)}{dt}=\upsilon_{t}\alpha E^{ash}\left( t \right)-\gamma_{2}J^{ash}\left( t \right)$$

$$\frac{dR^{ash}\left( t \right)}{dt}=\gamma_{1}I^{ash}\left( t \right)+\gamma_{2}J^{ash}\left( t \right)$$

where $\lambda_{t}^{ash}$: is the time-varying force of infection for individuals in group *a, s, h* at time *t*; *ψ_t_*: first-dose vaccination doses at time t*;* $\vartheta^{a}$: cumulative proportions of vaccines by age groups *a*; $\iota:$ 1-vaccine effectiveness (assuming leaky type); $\alpha$: rate at which individuals who acquired the infection become infectious (latent period)^−1^; *υ_t_*: proportion traced and isolated among exposed at time *t*; $\gamma_{1}$: effective rate of recovery among infectious individuals who are not traced and isolated = (effective infectious period)^−1^; $\gamma_{2}:$rate of recovery among infectious individuals who are traced and isolated = (self-isolation period)^−1^.

Additionally, we track the number of people with onset of symptoms ($O^{ash}$) and the cumulative number of people with symptoms that will be confirmed as mpox cases ($C^{ash})$, accounting for asymptomatic infections and underreporting ($\varepsilon)$and confirmation delays ($\eta)$.

$$\frac{dO^{ash}\left( t \right)}{dt}=\alpha E^{ash}\left( t \right)-{\eta O}^{ash}\left( t \right)$$

$$\frac{dC^{ash}\left( t \right)}{dt}=\varepsilon\eta O^{ash}\left( t \right)$$

#### *Force of infection and mixing patterns*

The force of infection ($\lambda_{t}^{ash})$was defined as the time-varying ($t$) per capita rate of mpox acquisition by age ($a)$, sexual activity ($s$), and HIV status-specific ($h)$ . It is a function of the time-varying sexual mixing matrix ($C_{ash,a^{'}s^{'}h^{'}}\left( t \right)$, reflecting an average number of sexual partnerships per-person per-day among group $ash$ with group $a's'h'$), transmission probability per effective contact (defined as a sexual partnership, $\beta$), and prevalence of people infectious with mpox among GBM available for sexual activities at time $t$ (i.e., not isolating). The force of infection is the following, where $N^{ash}$ is the size of the group $ash$.

$$\lambda_{t}^{ash}=\beta\sum_{a^{'}s^{'}h^{'}} C_{ash,a^{'}s^{'}h^{'}}\left( t \right)\frac{I^{a^{'}s^{'}h^{'}}\left( t \right)}{N^{a^{'}s^{'}h^{'}}-J^{a^{'}s^{'}h^{'}}\left( t \right)}$$

The time-varying mixing matrix $C_{ash,a^{'}s^{'}h^{'}}(t)$ was defined to reflect 5 factors: 1) changing numbers of non-isolating GBM and 2) changing contact rates during the mpox outbreak; and preferential mixing by 3) 5 age groups, 4) HIV status, and 5) 10 sexual activity groups.

*Mixing: group sizes and contact rates*

Numbers of non-isolating GBM in group $ash$ at time t were defined as $K^{ash}\left( t \right)= N^{ash}-J^{ash}\left( t \right)$.

Contact rates were defined as the number of sexual partners per-person per-day, including the rate ratio of the change in sexual partner numbers during the mpox outbreak (before first-dose vaccines were massively available on June 14^th^, 2022):

$$c^{ash}\left( t \right)=\left\{ \begin{aligned} c^{ash}, \mathrm{if} t<t_{\mathrm{mpo}x_{\mathrm{start}}} \mathrm{or} t\geq t_{\mathrm{mpo}x_{\mathrm{end}}} \\ c^{ash}\cdot RR, \mathrm{if} t_{\mathrm{mpo}x_{\mathrm{start}}}\leq t<t_{\mathrm{mpo}x_{\mathrm{end}}} \end{aligned} \right.$$

*Mixing: age and HIV status*

Mixing preferences by age and HIV status were informed by previous modelling of GBM in Montréal [27] (**Tables S3** and **S4**), while mixing preferences by sexual activity were specified parametrically with a single calibrated parameter $\omega$. All preferences were specified via odds ratios per [28], with iterative proportional fitting to maintain specified contact rates [29,30]. We defined a set of 15 (age) + 1 (HIV) odds ratios $\psi$, which were mapped to two symmetric matrices as follows:

$$\Psi_{aa^{'}}= \left[ \begin{matrix} \psi_{1} & \psi_{2} & \psi_{4} & \psi_{7} & \psi_{11} \\ \psi_{2} & \psi_{3} & \psi_{5} & \psi_{8} & \psi_{12} \\ \psi_{4} & \psi_{5} & \psi_{6} & \psi_{9} & \psi_{13} \\ \psi_{7} & \psi_{8} & \psi_{9} & \psi_{10} & \psi_{14} \\ \psi_{11} & \psi_{12} & \psi_{13} & \psi_{14} & \psi_{15} \end{matrix} \right], \Psi_{hh^{'}}=\left[ \begin{matrix} \psi_{0} & \cdot\\ \cdot& \psi_{0} \end{matrix} \right]$$

These matrices can be used to specify mixing by age and HIV status at the population-level (total number of contacts) via:

$$X_{ah}= \sum_{s} c^{ash}K^{ash}$$

$$X_{ah, a^{'}h^{'}}^{R}= \frac{X_{ah}X_{a^{'}h^{'}}}{\sum_{ah} X_{ah}}$$

$${X^{[0]}}_{ah, a^{'}h^{'}}= X_{ah, a^{'}h^{'}}^{R} \exp\left( \Psi_{aa^{'}}+\Psi_{hh^{'}} \right)$$

where $X_{ah}$ reflects the total numbers of contacts “offered” by group $ah$ (after summing over activity groups $s$), and $X_{ah, a^{'}h^{'}}^{R}$ reflects random (or “proportional”) mixing. The above definition of ${X^{\left[ 0 \right]}}_{ah, a^{'}h^{'}}$ changes the total numbers of contacts modelled for each group due to nonlinear effects of multiplication, but the original contact numbers can be recovered using iterative proportional fitting (denoted $\mathrm{IPF}(X)$) [29,30]:

$$X_{ah,a^{'}h^{'}}^{\left[ n+1 \right]}=X_{ah,a^{'}h^{'}}^{\left[ n \right]}\frac{X_{k}^{R}}{X_{k}^{\left[ n \right]}}, k=\left\{ \begin{aligned} ah, \mathrm{if}n is even \\ a^{'}h^{'}, \mathrm{if}n is odd \end{aligned} \right.$$

which typically converges to machine precision (${10}^{-12}$) within 5-50 iterations.

From the resulting population-level mixing matrix $X_{ah,a^{'}h^{'}}= X_{ah,a^{'}h^{'}}^{\left[ n \to\infty\right]}$, we can obtain probability-scale mixing matrices for age and HIV via:

$$p_{ah,a^{'}h^{'}}=\frac{X_{ah,a^{'}h^{'}}}{X_{ah}}, \hat{p}_{aa^{'}}=\frac{\sum_{hh^{'}} X_{ah,a^{'}h^{'}}}{\sum_{h} X_{ah}}, \hat{p}_{hh^{'}}=\frac{\sum_{aa^{'}} X_{ah,a^{'}h^{'}}}{\sum_{a} X_{ah}}$$

The matrices $\hat{p}_{aa^{'}}$ and $\hat{p}_{hh^{'}}$ should then match the data in **Tables S3** and **S4**, when using pre-mpox group sizes $N^{ash}$ and contact rates $c^{ash}$ to compute $X$. Thus, we estimated $\psi$ by minimizing the mean absolute differences between $p_{aa^{'}}$ and $p_{hh^{'}}$ from the data (**Tables S3** and **S4**) and $\hat{p}_{aa^{'}}$ and $\hat{p}_{hh^{'}}$ from the parametric model for mixing described above. The advantage of this odds-based approach is that mixing matrices can remain both “balanced” and reflective of empiric mixing preferences despite changes to effective group sizes $K^{ash}(t)$ and contact rates $c^{ash}(t)$ during the simulated epidemic.

*Mixing: sexual activity*

We used a similar approach as above to define the time-varying mixing matrix further stratified by sexual activity: $X_{ash,a^{'}s^{'}h^{'}}(t)$. First, we defined the total numbers of contacts “offered” by group $ash$ to group $a^{'}h^{'}$ as:

$$X_{ash\left[ a^{'}h^{'} \right]}\left( t \right)= K^{ash}\left( t \right) c^{ash}\left( t \right) p_{ah,a^{'}h^{'}}\left( t \right)$$

which incorporates mixing preference by age and HIV status. Then we specify increased odds of mixing by sexual activity group (conditional on age and HIV status) via a matrix $\Psi_{ss^{'}}$ applied to the random mixing matrix, and adjusted with $\mathrm{IPF}$:

$$X_{ash,a^{'}s^{'}h^{'}}^{R}\left( t \right)=\frac{X_{ash\left[ a^{'}h^{'} \right]}X_{a^{'}s^{'}h^{'}\left[ ah \right]}}{\sum_{ash} X_{ash\left[ a^{'}h^{'} \right]}}$$

$$X_{ash,a^{'}s^{'}h^{'}}\left( t \right)= \mathrm{IPF}\left( X_{ash,a^{'}s^{'}h^{'}}^{R}\exp\Psi_{{ss}^{'}} \right)$$

In this case, we had no data to inform the odds matrix $\Psi_{ss^{'}}$. So, we assumed a gaussian “fuzzy diagonal” with fixed mean $\mu=0$ and standard deviation $\sigma=2$ (95% probability mass within ±5 activity groups), whose overall magnitude was scaled by the calibrated degree of assortativity parameter, $\omega\in(0, 100)$:

$$\Psi_{ss^{'}}=\omega\cdot\mathrm{norm}\left( \left| s-s^{'} \right|,\mu,\sigma\right)\cdot\sigma$$

Finally, we converted the resulting population-level mixing matrix $X$ to the per-person scale $C$ for use in the force of infection equation via:

$$C_{ash,a^{'}s^{'}h^{'}}\left( t \right)=\frac{X_{ash,a^{'}s^{'}h^{'}}}{K^{ash}}$$

#### *Model parameterization*

*Population sizes*

Using previous estimates of the GBM population size in each city [1,31,32], we modeled 54,000, 78,000, and 26,100 sexually active GBM in Montréal, Toronto, and Vancouver, respectively. The prevalence of HIV among GBM was estimated from the *Engage* baseline visit, accounting for the RDS survey design by incorporating the RDS-II weights [33]. The age distribution was informed by the *Canadian 2021 Census of Population* [32]. To model only sexual active GBM, we adjusted the size of the population aged ≥60 years old in our model, as sexual activity tends to decline at older ages [34]. The *Institut de la statistique du Québec* estimated 35.6% of men ≥65 years old were sexually active in 2020-2021 [34], which is similar to estimates from a 2022 UK study of GBM [4]. We assumed this proportion applied to GBM and was similar across Montréal, Toronto, and Vancouver.

*Sexual activity groups*

To describe the contact rates among GBM, we obtained the distribution of sexual partner numbers during 2022 prior to the first reported mpox case in Canada (May 19^th^, 2022). Specifically, we leveraged our previous results from a Bayesian negative binomial regression model where we described the distributions of sexual partner numbers for participants in each combination of age group and HIV status [1]. We chose to use the fitted, as opposed to the empirical distribution, to account for *Engage*’s RDS study design and loss to follow-up. This was achieved by incorporating RDS-II and inverse probability of censoring weights via post-stratification. Then, we partitioned each age-HIV status combination into 10 sexual activity groups based on 60%, 90%, 95%, 96%, 97%, 98%, 99%, 99.5%, and 99.8% percentiles, which effectively described the heterogeneity in GBM partner numbers and captured well the heavy-tailed distributions [35]. We converted the P6M contact rates to daily rates. This procedure was repeated for each of the three cities independently to obtain city-specific contact rates for each combination of age group, sexual activity group, and HIV status.

*Modeled change in sexual partner numbers*

We used the rate ratio from the analysis of sexual partner numbers to inform the prior for the RR parameter in the model. The RR parameter is calibrated to account for a potential reduction in the contact rate during the period of mpox-driven behaviour changes (May 19^th^-August 14^th^, 2022) and capped it at 1, as increases in sexual activity during mpox are qualitatively unlikely. Furthermore, we allowed the RR to vary based on whether the sexual activity group’s contact rate was defined by ≤7 or >7 partners, as informed by the empirical analyses.

*Contact tracing/isolation*

We assumed that since the first reported case in each city, 20% of contacts were identified through contact tracing and were advised to self-isolated, based on data from *Direction régionale de santé publique de* *Montréal* [18]. We assumed that a similar proportion of cases would be traced in the two other cities [8,18].

Compartmental models, such as ours, cannot accurately represent contact tracing activities. Hence, we adjusted the 20% of contact traced for several factors. First, we posited that traced and untraced contacts have the same probability of having acquired mpox. Second, we assumed that only contacts of reported cases can be traced. The city-specific reporting fractions were informed by the estimates from our previous study: 82% for Montréal, 86% for Toronto, and 77% for Vancouver [1]. Third, we conservatively assumed that sexual contacts were exposed 4 days before a contact was traced: 2 days to confirm a reported cases and 2 days to perform the contact tracing. Fourth, we accounted for the fraction of contacts that would still be in the latent period 4 days after being infected. Based on previous studies [13–17], GBM stay on average 5.1 days in the exposed state and the underlying assumption of compartmental models is that the time spent in a compartment is exponentially distributed. The fraction of contacts traced before being infectious is therefore estimated as the cumulative proportion in the exposed compartment after 4 days (2 days for contact tracing and 2 days for reporting delay), as following:

$$Fraction of contacts traced before infectiousness=P\left( T_{exposed}\geq4 \right)=46\%$$

where $T_{exposed} \sim Exp(\frac{1}{5.1})$.

Hence, the fraction of exposed cases that are traced and isolated is 7.5% in Montréal (20%×82%×46%), 7.9% in Toronto (20%×86%×46%), and 7.0% in Vancouver (20%×77%×46%).

*Vaccinations*

The *Direction régionale de santé publique de* *Montréal, Public Health Ontario*, and the *British Columbia Centre for Disease Control* published data on weekly numbers of first-dose vaccination administered [9,18,19]. We did not model second-dose vaccinations as the great majority of second-doses were administered during Fall 2022, at a time where the outbreaks had been controlled [9,18,19]. All first-dose vaccines were assumed to have been offered as pre-exposure prophylaxis in the model (i.e., prior to a potential exposure). This is a reasonable assumption given only a small proportion of all doses administered were PEP (<3% in Ontario) [19], and those offered PEP were also likely to be notified through contact tracing and isolated, which was accounted for by the isolated compartment. We utilized city-level vaccination doses for Montréal and Vancouver, and provincial administered doses for Toronto (assumed all allocated to the city). We assumed that all vaccines were administered to GBM, as only sexually active GBM, sex workers, and workers on sex-on-premises venues were eligible for the PrEP vaccination in Canada [36]. Vaccination was attributed to each age group according to the proportion of vaccines received by each age group from immunization reports (***Table S5***) [8,19]. In Vancouver, the information was unavailable, and we therefore estimated this distribution by averaging the vaccine’s age distribution from the other two cities. Among each age group, vaccines were allocated proportionally to combinations of sexual-activity-HIV status groups according to their respective size. Vaccine effectiveness of single-dose mpox vaccines were informed by our meta-analysis of 7 recent studies (***Figure S1***) [3,20–25].

*Seeding of the epidemics*

We seeded the mpox epidemics according to the predominant characteristics of people with confirmed mpox from early case investigations [37]. Specifically, we imported cases among the 5% highest activity groups and among the 16-29, 30-39, and 40-49 age groups. Cases were allocated proportionally to the relative size of the groups and distributed equally to the exposed and infectious compartments. The number of imported cases were calibrated to reflect the uncertainty in the parameter.

As the outbreak first took place in Montréal, followed shortly by Toronto, and last occurred in Vancouver [9,10,18], we assumed a duration of 21 days from case importation to first reported cases in Montréal and Toronto, and 10 days for Vancouver.

#### *Model calibration*

*Confirmed mpox cases*

I utilized data on mpox reported to public health agencies (i.e., “reported cases”), including daily confirmed and probable mpox cases. A confirmed mpox case is an individual detected of mpox virus DNA by a nuclear acid amplification test. A probable case is a person detected of a virus from the genus *Orthopoxvirus*, a suspect case with substantial exposure to a confirmed case in the 21 days before symptom onset, or a suspect case who was male and had sex with a man in the 21 days before symptom onset [18].

First, provincial daily cases for Québec, Ontario, and British Columbia were publicly available from the website of the *Public Health Agency of Canada* [37]. Only the total (confirmed and probable) cases counts were available for Québec.

I then used weekly-varying city fractions of provincial cases to estimate the confirmed cases in each city. For Montréal, the *Direction régionale de santé publique de Montréal* published the weekly-varying fractions of total provincial cases that were from the city [18]. For Toronto, the weekly-varying number of total city cases was available from a publication by the *Public Health Ontario* [38]. Using this data, we were able to estimate weekly-varying total city cases over confirmed provincial cases. Finally, the weekly fractions of confirmed provincial cases that were from the city was available from a surveillance report by the *British Columbia Centre for Disease Control* [9].

To estimate the daily confirmed cases in Montréal, I leveraged the weekly fractions of total reported cases that were confirmed in the city [8], and multiplied this quantity with the daily provincial total cases and the weekly-varying fractions of total provincial cases that were from the city. For Toronto, the fraction of probable cases in Ontario that were confirmed was 98% as of June 2023 [38], and we estimated the confirmed city cases in a similar fashion as in Montréal. Finally, the daily confirmed cases in Vancouver were estimated using the product of the daily provincial cases and fractions of confirmed provincial cases that were confirmed in the city.

*Calibration algorithm*

We used a Bayesian framework to obtain posterior distributions of parameters and outcomes [49]. First, the model parameters were calibrated to daily (*d*) incidence of observed mpox cases ($C_{d}^{observed})$in the cities using a negative binomial likelihood with mean equal to the daily incidence of modeled mpox cases ($C_{d}^{modeled})$ and overdispersion parameter equal to 0.1.

$$C_{d}^{observed}\sim NB\left( C_{d}^{modeled}, 0.1 \right)$$

$$C_{d}^{modeled}=\sum_{s=1}^{20} \frac{dC^{s}\left( t \right)}{dt}\left. \right|_{t=d}$$

We then performed optimization, using the Broyden-Fletcher-Goldfarb-Shanno (BFGS) algorithm, to obtain the posterior modes (maximum a posteriori estimates) of the parameters. We utilized a sampling importance resampling (SIR) algorithm with 5,000 parameter sets from the proposal distribution estimated from the optimization routine (multivariate *t* distribution with 2 degrees of freedom).

The three city were calibrated together and five parameters were estimated: 1) the number of imported cases ($\tau$), 2) transmission probability per effective contact (defined as a sexual partnership, $\beta$), 3) duration of the effective infectious period (defined as time from infectivity onset to sexual abstinence or recovery, ${\gamma_{1}}^{-1}$), 4) the degree of assortativity by risk groups (i.e., mixing parameter, $\omega$), and 5) the RR of contact rate during periods of mpox-driven behaviour changes (*RR*). All parameters, except the number of imported cases and the degree of assortativity, were assumed constant across the cities.

The prior distributions for the duration of infectiousness and transmission probability per effective contact were derived from a previous modelling study [1]. Prior distributions for the number of imported cases were informed by the relative size of the mpox outbreak in each city [37]. The prior distribution for the RR was informed by the change in sexual partner numbers analysis: the prior of RR among the ≤7 sexual activity level ($RR_{\leq7 partners}$) was capped between the lower bound of the 95% CrI (0.47) and 1. Since GBM of >7 sexual activity level likely have a greater decrease in partner numbers during the mpox outbreak (i.e., a lower RR), we constraint $RR_{>7 partners}$ to be lower than $RR_{\leq7 partners}$. This was achieved by adding an $RR_{\mathrm{multiplier}}=\frac{RR_{>7 partners}}{RR_{\leq7 partners}}$ smaller than 1. Finally, the prior for the assortativity parameter was capped under 100 and was determined by manual fitting to the observed epidemics. The priors are the following:

$$\gamma_{1}^{-1}\sim\left( 3+{logit}^{-1}\left( N\left( logit\left( \frac{5 - 3}{15-3} \right), 1 \right) \right)\times\left( 15-3 \right) \right)$$

$$\beta\sim{logit}^{-1}\left( N\left( logit\left( 0.87 \right), 1 \right) \right)$$

$$RR_{\leq7 partners}\sim0.47+{logit}^{-1}\left( N\left( logit\left( \frac{0.80 - 0.47}{1-0.47} \right), 1.5 \right) \right)\times(1-0.47)$$

$$RR_{\mathrm{multiplier}}\sim{0.70+logit}^{-1}\left( N\left( logit\left( \frac{0.67}{0.80} \right), 5 \right) \right)\times(1-0.70)$$

$$\tau_{t_{0}}^{c}\sim\left\{ \begin{matrix} 2+{logit}^{-1}\left( N\left( logit\left( 0.5 \right), 0.5 \right) \right)\times\left( 8-2 \right);if c= Montréal \\ 3+{logit}^{-1}\left( N\left( logit\left( 0.5 \right), 0.5 \right) \right)\times\left( 8-3 \right);if c= \mathrm{Toronto} \\ 1+{logit}^{-1}\left( N\left( logit\left( 0.5 \right), 0.5 \right) \right)\times\left( 6-1 \right);if c= \mathrm{Vancouver} \end{matrix} \right.$$

$$\omega\sim{logit}^{-1}\left( N\left( logit\left( \frac{5}{100} \right), 1 \right) \right)\times100$$

### Supplementary Tables

#### Table S1. Variables used to empirically estimate changes in numbers of sexual partners during the mpox outbreak using data from the *Engage Cohort Study*.

| **Notation** | **Definition** | **Domain** | **Question from the *Engage Cohort Study*** |
| --- | --- | --- | --- |
| ***y*** | Number of all-type sexual partners in the past 6 months (P6M) reported at the 2022 visit | $[0,1000]$ | 5.5 During the PAST 6 MONTHS, with how many guys have you had any kind of sex (anal, oral, mutual masturbation, rimming, frontal/vaginal, etc.)?  _____ guys |
| ***Age*** | Age at time of the 2022 visit | 16-29, 30-39, 40-49, 50-59, ≥60 years | 1.3 What is your age (i.e., how old are you)? |
| ***Relationship status history*** | Relationship status reported at the latest visit before 2022 | single, exclusive relationship, open relationship, unclear | 2.18 Do you currently have a relationship with a main partner? No \| Yes  2.23 What discussions have you and your main partner had with each other in terms of only having sex with each other?  We haven’t explicitly discussed only having sex with each other or not \| We have discussed only having sex with each other, but have not agreed to anything \| We have discussed only having sex with each other and agreed to only have sex with each other \| We agreed to have other sex partners, but only ones we share (we only play together) \| We agreed to have other sex partners, some of whom we share and others whom we see separately (we play together and separately) \| We agreed to have other sex partners whom we only see separately (we only play separately) \| We agreed to another arrangement. Please describe: __ \| No main relationship partner |
| ***HIV status*** | HIV status at time of the 2022 visit | Seropositive, seronegative | Derived by *Engage Cohort Study* from participant laboratory-tested and self-report HIV status (determined using 4^th^ generation testing with a confirmatory assay; or self-reported if testing data unavailable) |
| ***Calendar month*** | Calendar month of visit during 2022 | {1, 2, …, 12}; continuous | - |
| ***Sexual partnership history*** | Number of all-type sexual partners in the past 6 months reported at the latest visit before 2022 | ≤ 7, >7 | 5.5 During the PAST 6 MONTHS, with how many guys have you had any kind of sex (anal, oral, mutual masturbation, rimming, frontal/vaginal, etc.)?  _____ guys |

**Table S2.** Parameter values used for the mpox dynamic transmission model in Montréal, Toronto, and Vancouver (2022).

| **Parameters** | **Unit** | **Symbol** | **Value** | **95% CrI of prior density*** | **Sources** |
| --- | --- | --- | --- | --- | --- |
| ***Outbreak parameters*** | | |  |  |  |
| **Number of imported cases^‡^** | cases | $\tau$ | — | Montréal: (3.6, 6.3)  Toronto: (4.4, 6.6)  Vancouver: (2.4, 4.6) | Calibrated[37] |
| **Reporting delay** | day | $\eta^{-1}$ | 2 | — | Assumed |
| **Reporting fraction** | % | $\varepsilon$ | Montréal: 82%  Toronto: 86%  Vancouver: 77% | — | [1] |
| ***Natural history parameters*** | | |  |  |  |
| **Incubation period^†^** | day |  | 7.1 | — | [14–17] |
| **Latent period** | day | $\alpha^{-1}$ | 5.1 | — | [13] |
| **Effective infectious period among GBM not traced and isolating** | day | ${\gamma_{1}}^{-1}$ | — | (4.7, 13.7) | Calibrated [1,17,39] |
| **Self-isolation period** | day | $\gamma_{2}^{-1}$ | 14 | — | [40–42] |
| **Risk of transmission per effective contact** | % | $\beta$ | — | (49%, 98%) | Calibrated [1,43] |
| ***Public health intervention parameters*** | | |  |  |  |
| **Percentage traced and isolated among the exposed** | % | $v_{t}$ | Montréal: 7.5%  Toronto:7.9%  Vancouver: 7.0% | — | [8,18] |
| **First-dose vaccination doses** | doses | $\psi_{t}$ | Time-varying | — | [9,18,19] |
| **Percentage vaccinations received by age groups** | % | $\vartheta$ | ***See Table S5*** | — | [8,19] |
| **Vaccine effectiveness of the first dose**^†^ | % | $\iota$ | 51.5% | — | [3,20–25] |
| ***Sexual behaviour parameters*** | | |  |  |  |
| **GBM population size** | persons |  | Montréal: 54,000  Toronto: 78,000  Vancouver: 26,100 | — | [1,31,32] |
| **Mixing parameter** |  | $\omega$ | — | (0.75, 27.40) | Calibrated [1] |
| **Reduction in partner change rate during mpox** |  | *RR* | — | ≤7 sexual partners (P6M): (0.51, 0.99)  >7 sexual partners (P6M): (0.40, 0.97) | Calibrated |
| *GBM,* gay, bisexual, and other men who have sex with men; *CrI*, credible interval. * For parameters to be calibrated, the 95% CrI from the prior density distribution (described in ***Supplementary Methods***) was shown. † Values estimated from a meta-analysis of studies (***Figure S1***). | | | | | |

#### **Table S3: Age mixing matrix (**$\boldsymbol{p}_{\boldsymbol{a}\boldsymbol{a}^{\boldsymbol{'}}}$**).** The mixing matrix represents the proportion of GBM in each age group who reported that the age of their last sexual partner fell within the corresponding age group in each city. The data was derived from the age mixing matrix from Milwid et al. (2022) [27] and the age distribution from the *Engage Cohort Study (2017-2023)*.

| Montréal | Partner’s age group | | | | | |
| --- | --- | --- | --- | --- | --- | --- |
| GBM’s age group |  | *16-29* | *30-39* | *40-49* | *50-59* | *>60* |
|  | *16-29* | 0.716 | 0.181 | 0.079 | 0.018 | 0.005 |
|  | *30-39* | 0.284 | 0.37 | 0.292 | 0.036 | 0.017 |
|  | *40-49* | 0.161 | 0.369 | 0.361 | 0.088 | 0.022 |
|  | *50-59* | 0.144 | 0.281 | 0.387 | 0.161 | 0.027 |
|  | *>60* | 0.497 | 0.141 | 0.13 | 0.175 | 0.056 |
| Toronto | **Partner’s age group** | | | | | |
| GBM’s age group |  | *16-29* | *30-39* | *40-49* | *50-59* | *>60* |
|  | *16-29* | 0.715 | 0.182 | 0.079 | 0.019 | 0.005 |
|  | *30-39* | 0.288 | 0.369 | 0.291 | 0.036 | 0.017 |
|  | *40-49* | 0.161 | 0.368 | 0.361 | 0.088 | 0.022 |
|  | *50-59* | 0.144 | 0.281 | 0.388 | 0.161 | 0.027 |
|  | *>60* | 0.492 | 0.144 | 0.133 | 0.174 | 0.057 |
| Vancouver | **Partner’s age group** | | | | | |
| GBM’s age group |  | *16-29* | *30-39* | *40-49* | *50-59* | *>60* |
|  | *16-29* | 0.706 | 0.187 | 0.082 | 0.019 | 0.005 |
|  | *30-39* | 0.287 | 0.369 | 0.291 | 0.036 | 0.017 |
|  | *40-49* | 0.161 | 0.369 | 0.361 | 0.088 | 0.022 |
|  | *50-59* | 0.144 | 0.281 | 0.388 | 0.16 | 0.027 |
|  | *>60* | 0.497 | 0.141 | 0.131 | 0.174 | 0.056 |

Table S4: Mixing matrix by HIV status ($p_{hh^{'}}$) for Montréal. The mixing matrix represents the proportion of partnerships among GBM of a given HIV status with partners living or not living with HIV. “HIV- / unknown” represents GBM who received a negative test result or do not know their HIV status. The matrix was taken from Milwid et al. (2022) [27].

|  | Partner’s HIV status | | |
| --- | --- | --- | --- |
| GBM’s HIV status |  | *HIV- / unknown* | *HIV+* |
|  | *HIV- / unknown* | 0.924 | 0.076 |
|  | *HIV+* | 0.660 | 0.340 |

Table S5: Proportion of mpox vaccines received by age groups in Montréal and Toronto. The data was from the *Direction régionale de santé publique de* *Montréal* and *Public Health Ontario* [8,19]. Proportions for 50-59 and 60+ age groups were not available, and vaccines were assumed to be distributed equally between the two groups.

| **Age** | **Proportion of mpox vaccines** | |
| --- | --- | --- |
|  | **Montréal** | **Toronto** |
| 16-29 | 21.5% | 22.1% |
| 30-39 | 26.5% | 33.4% |
| 40-49 | 20.0% | 18.2% |
| 50-59 | 16.0% | 13.2% |
| 60+ | 16.0% | 13.2% |

Table S6: Unadjusted and RDS-II adjusted characteristics of *Engage Cohort Study* visits in 2022. Unadjusted and RDS-II adjusted estimates of percent coverage of the period of mpox-driven behaviour changes, number of sexual partners in the past six months, and covariates.

|  | **Visit during the rest of 2022** | | | | **Visit during the period of** **mpox-driven behaviour changes** | | | | | | **Overall** | | | | |
| --- | --- | --- | --- | --- | --- | --- | --- | --- | --- | --- | --- | --- | --- | --- | --- |
|  | $\boldsymbol{N}$***/mean*** | **(%)** | **RDS-II weighted % (95% CI)** | | $\boldsymbol{N}$***/mean*** | | **(%)** | | **RDS-II weighted % (95% CI)** | | $\boldsymbol{N}$***/mean*** | **(%)** | | **RDS-II weighted % (95% CI)** | |
| ***Number of participant visits*** | 1533 |  |  |  | 424 |  | |  | |  | 1957 |  |  | |  |
| ***Number of participants*** | 1078 |  |  |  | 367 |  | |  | |  | 1445 |  |  | |  |
| ***ESS of participants*** | 721 |  |  |  | 174 |  | |  | |  | 547 |  |  | |  |
| ***Percent coverage (%) of the period of*** ***mpox-driven behaviour changes (mean)*** | | | | | | | | | | |  | | | | |
|  | 0.0% | (SD0%) | 0% | (0–0%) | 19.5% | (SD13.4%) | | 19.5% | | (16.3–22.8%) | 4.2% | (SD10.2%) | 3.8% | | (3.1–4.5%) |
| ***Months since 2022-01-01 (mean)*** | | | | |  | | | | | |  | | | | |
|  | 5.1 | (SD3.5) | 5.3 | (4.7–6) | 6.2 | (SD0.9) | | 6.2 | | (5.3–7.1) | 5.4 | (SD3.2) | 5.5 | | (4.9–6.1) |
| ***Age group*** | | | | |  | | | | | |  | | | | |
| *16-29* | 217 | (14%) | 18% | (16–20%) | 68 | (16%) | | 24% | | (19–28%) | 285 | (15%) | 19% | | (17–21%) |
| *30-39* | 613 | (40%) | 40% | (37–42%) | 196 | (46%) | | 39% | | (34–44%) | 809 | (41%) | 40% | | (37–42%) |
| *40-49* | 273 | (18%) | 13% | (11–14%) | 70 | (17%) | | 16% | | (12–20%) | 343 | (18%) | 13% | | (12–15%) |
| *50-59* | 203 | (13%) | 11% | (9–13%) | 45 | (11%) | | 9% | | (6–12%) | 248 | (13%) | 10% | | (9–12%) |
| *60+* | 227 | (15%) | 18% | (16–20%) | 45 | (11%) | | 13% | | (10–17%) | 272 | (14%) | 17% | | (16–19%) |
| ***Relationship status history*** | | | | |  | | | | | |  | | | | |
| *Single* | 818 | (53%) | 54% | (51–57%) | 222 | (52%) | | 51% | | (46–57%) | 1040 | (53%) | 54% | | (51–56%) |
| *Open* | 426 | (28%) | 19% | (17–21%) | 136 | (32%) | | 29% | | (24–34%) | 562 | (29%) | 21% | | (19–23%) |
| *Exclusive* | 205 | (13%) | 20% | (18–22%) | 44 | (10%) | | 14% | | (10–17%) | 249 | (13%) | 19% | | (17–21%) |
| *Unclear* | 84 | (5%) | 6% | (5–8%) | 22 | (5%) | | 6% | | (3–8%) | 106 | (5%) | 6% | | (5–7%) |
| ***HIV status**** | | | | |  | | | | | |  | | | | |
| *Seropositive* | 312 | (20%) | 18% | (16–20%) | 52 | (12%) | | 9% | | (6–12%) | 364 | (19%) | 16% | | (14–18%) |
| *Seronegative / unknown* | 1221 | (80%) | 82% | (80–84%) | 372 | (88%) | | 91% | | (88–94%) | 1593 | (81%) | 84% | | (82–86%) |
| ***Sexual partner numbers at the latest visit before 2022*** | | | | |  | | | | | |  | | | | |
| *≤ 7 sexual partners* | 1211 | (79%) | 88% | (86–89%) | 324 | (76%) | | 83% | | (79–87%) | 1535 | (78%) | 87% | | (85–88%) |
| *> 7 sexual partners* | 322 | (21%) | 12% | (11–14%) | 100 | (24%) | | 17% | | (13–21%) | 422 | (22%) | 13% | | (12–15%) |
| ***Number of sexual partners in the past 6 months (mean)*** | | | | |  | | | | | |  | | | | |
|  | 7.1 | (SD14.1) | 4.6 | (4–5.3) | 7.4 | (SD14.4) | | 5.3 | | (3.8–6.8) | 7.1 | (SD14.2) | 4.8 | | (4.2–5.4) |
| *N:* number of visits (unless specified in the first column); *CI*, confidence interval; *ESS*, effective sample size (of participants) is the size of a simple random sample that would produce the same variance as the RDS-II design and was estimated using *survey* package in R; *RDS*, respondent driven sampling; *SD*, standard deviation. ^*^HIV status was ascertained from 4^th^ generation laboratory testing with a confirmatory assay. Self-reported status was used if the test result was unknown (number of visits=52). RDS-II weights are inversely proportional to participants’ social network size. | | | | | | | | | | | | | | | |

#### **Table S7**. Association between number of sexual partners in the past 6 months and covariates among *Engage Cohort Study* participants during the period of mpox-driven potential behaviour changes (May 19^th^ –August 14^th^, 2022).

|  | **RR (95% CrI)** |
| --- | --- |
| ***Period of mpox-driven behaviour changes*** | 0.80 (0.47, 1.36) |
| ***Months since January 1^st^, 2022*** | 1.01 (1, 1.03) |
| ***Age group*** |  |
| ***16-29*** | Reference |
| ***30-39*** | 0.91 (0.77, 1.09) |
| ***40-49*** | 0.85 (0.69, 1.04) |
| ***50-59*** | 0.62 (0.5, 0.78) |
| ***≥60*** | 0.51 (0.41, 0.64) |
| ***Relationship status history*** |  |
| ***Single*** | Reference |
| ***Open*** | 1.44 (1.27, 1.64) |
| ***Exclusive*** | 0.74 (0.61, 0.91) |
| ***Unclear*** | 0.82 (0.62, 1.09) |
| ***HIV seropositive**** | 1.13 (0.97, 1.31) |
| ***Sexual partner numbers at the latest visit before 2022*** | |
| ***≤ 7 sexual partners*** | Reference |
| ***> 7 sexual partners*** | 5.61 (4.9, 6.45) |
| ***Period of mpox-driven behaviour changes by sexual partners interaction*** | 0.83 (0.32, 2.15) |
| ***1 / overdispersion parameter*** | 3.7 (3.02, 4.47) |
| Table presents the mean and 95% credible interval of a negative binomial regression model with a random intercept for each participant estimated using partial pooling. The percent coverage of the period of mpox-driven behaviour changes and calendar month variables are centred.  *CrI*, credible interval; *RR*, rate ratio.  * HIV status was determined based on 4^th^ generation testing with a confirmatory assay. If the laboratory test result was unknown, self-reported status was used. | |

Table S8. Effect of the mpox epidemic on the number of sexual partners and visits to sex-on-premises venues. Sensitivity analyses using various sexual activity level groupings, endpoints for the mpox outbreak period, or using visit to bathhouses and/or sex clubs at least once in the P6M or attendance of group sex events at least once in the P6M as the outcome.

| **Analysis** | **Partner numbers at the latest visit before 2022** | | $\boldsymbol{N}$ | | **RR (95% CrI)** | |
| --- | --- | --- | --- | --- | --- | --- |
|  |  | | *period of potential behavior change* | *rest of 2022* | | |
| ***Using the 2022-08-14 endpoint*** | | | | | | |
|  | | ***≤ 3 sexual partners*** | 251 | 930 | 0.91 (0.48, 1.67) | |
|  | | ***> 3 sexual partners*** | 173 | 603 | 0.75 (0.31, 1.83) | |
|  | | ***≤ 5 sexual partners*** | 299 | 1112 | 0.85 (0.49, 1.49) | |
|  |  | ***> 5 sexual partners*** | 125 | 421 | 0.64 (0.29, 1.41) | |
| ***(main analysis)*** | | ***≤ 7 sexual partners*** | 324 | 1211 | 0.80 (0.47, 1.36) | |
|  |  | ***> 7 sexual partners*** | 100 | 322 | 0.67 (0.31, 1.43) | |
| ***Using the 2022-07-14 endpoint*** | | | | | | |
|  | | ***≤ 3 sexual partners*** | 197 | 984 | | 1.48 (0.55, 4.06) |
|  | | ***> 3 sexual partners*** | 132 | 644 | | 0.86 (0.20, 3.66) |
|  | | ***≤ 5 sexual partners*** | 235 | 1176 | | 0.99 (0.39, 2.54) |
|  | | ***> 5 sexual partners*** | 94 | 452 | | 0.92 (0.25, 3.41) |
|  | | ***≤ 7 sexual partners*** | 249 | 1286 | | 1.03 (0.42, 2.57) |
|  |  | ***> 7 sexual partners*** | 80 | 342 | | 0.54 (0.15, 1.98) |
| ***Using* visit to bathhouses and/or sex clubs as the outcome** | | | | | | |
|  | |  | 424 | 1533 | | 0.26 (0.03, 1.89) |
| ***Using* attendance of group sex events as the outcome** | | | | | | |
|  | |  | 424 | 1533 | | 2.42 (0.25, 23.8) |
| Models adjusted for month of the visit (1, 2,…,12; continuous), age (16-29, 30-39, 40-49, 50-59, ≥60 years), relationship status history (single, exclusive relationship, open relationship, unclear), and HIV status (seropositive, seronegative). $N$, number of visits; *CrI*, credible interval; *RR*, rate ratio of the percent coverage of the period of period of mpox-driven behaviour changes to the number of sexual partners in the past 6 months. | | | | | | |

#### Table S9. Calibrated parameters for the dynamic model of mpox transmission in Montréal, Toronto, and Vancouver.

| **Parameters** | Unit | $Symbol$ | Prior median (95% CrI) | Posterior median (95% CrI) |
| --- | --- | --- | --- | --- |
| **Effective infectious period among GBM not isolating** | day | ${\gamma_{1}}^{-1}$ | 9.50 (4.71,13.72) | 5.87 (4.83, 7.15) |
| **Risk of transmission per effective contact** | % | $\beta$ | 87 (49, 98) | 86 (63, 95) |
| **Change in sexual partner numbers** |  | *RR* | ≤7 sexual activity level:  0.81 (0.51, 0.99)  >7 sexual activity level:  0.63 (0.30, 0.91) | ≤7 sexual activity level:  0.94 (0.80, 0.99)  >7 sexual activity level:  0.94 (0.70, 0.99) |
|  |  |  | Montréal: 5.00 (3.61, 6.34)  Toronto: 5.50 (4.37, 6.61)  Vancouver: 3.50 (2.37, 4.64) | Montréal: 4.98 (3.53, 6.17)  Toronto: 5.44 (4.26, 6.34)  Vancouver: 3.40 (2.43, 4.46) |
| **Number of imported cases^‡^** | cases | $\tau$ |  |  |
| **Mixing parameter** |  | $\omega$ | 5.06 (0.75, 27.40) | Montréal: 6.17 (3.85, 8.35)  Toronto: 6.92 (2.87, 14.58)  Vancouver: 10.95 (7.40, 18.81) |
| *CrI*, credible interval.  ‡ Values shown are city-specific parameter estimate and 95% CrI’s. | | | | |

#### Table S10. Median averted fraction (AF) of mpox cases due to interventions that led to change in numbers of sexual partners, contact tracing/isolation, and first-dose vaccination in Montréal, Toronto, and Vancouver (Canada). The estimates from the main scenario are presented, as well as several sensitivity analyses.

| **Analyses** | **City** | **Averted Fraction (95% CrI)** | | | | | | |
| --- | --- | --- | --- | --- | --- | --- | --- | --- |
|  |  | Change in sexual partner numbers | Contact tracing/isolation | Vaccination | All three combined | Change in sexual partner numbers and contact tracing/isolation | Change in sexual partner numbers and vaccination | Contact tracing/isolation and vaccination |
| *Main (as reported in manuscript)* | Montréal | 15% (3%-34%) | 14% (12%-21%) | 21% (16%-33%) | 48% (35%-66%) | 29% (17%-48%) | 36% (23%-55%) | 33% (27%-49%) |
|  | Toronto | 11% (2%-27%) | 14% (12%-22%) | 22% (16%-41%) | 46% (34%-67%) | 25% (16%-40%) | 36% (22%-56%) | 33% (26%-57%) |
|  | Vancouver | 10% (2%-22%) | 14% (12%-16%) | 39% (35%-48%) | 58% (52%-69%) | 23% (16%-33%) | 50% (43%-60%) | 49% (44%-59%) |
| *Informative prior for RR*^‡^ | Montréal | 49% (44%-53%) | 11% (10%-13%) | 15% (13%-20%) | 67% (59%-77%) | 56% (51%-63%) | 61% (54%-70%) | 26% (22%-32%) |
|  | Toronto | 39% (31%-42%) | 12% (11%-14%) | 17% (15%-26%) | 62% (59%-71%) | 46% (42%-49%) | 56% (53%-64%) | 27% (25%-38%) |
|  | Vancouver | 29% (26%-32%) | 12% (11%-13%) | 34% (32%-38%) | 70% (67%-73%) | 38% (35%-42%) | 65% (62%-67%) | 42% (40%-46%) |
| *10% of contract traced (instead of 20%)* | Montréal | 16% (3%-35%) | 7% (6%-11%) | 22% (16%-33%) | 44% (30%-61%) | 24% (10%-41%) | 38% (24%-55%) | 28% (22%-42%) |
|  | Toronto | 13% (2%-28%) | 7% (6%-11%) | 22% (16%-41%) | 42% (29%-62%) | 20% (10%-34%) | 37% (23%-58%) | 27% (21%-50%) |
|  | Vancouver | 11% (2%-23%) | 7% (6%-9%) | 40% (35%-50%) | 56% (48%-66%) | 18% (9%-29%) | 51% (44%-62%) | 45% (40%-55%) |
| *15% of contact traced (instead of 20%)* | Montréal | 15% (3%-36%) | 11% (9%-17%) | 21% (16%-34%) | 46% (33%-63%) | 26% (14%-46%) | 37% (24%-55%) | 31% (24%-47%) |
|  | Toronto | 12% (2%-28%) | 10% (9%-16%) | 22% (16%-41%) | 45% (31%-65%) | 22% (13%-37%) | 36% (23%-57%) | 30% (24%-53%) |
|  | Vancouver | 10% (2%-23%) | 10% (9%-12%) | 40% (35%-48%) | 57% (50%-67%) | 20% (13%-31%) | 50% (43%-62%) | 47% (42%-56%) |
| *Vaccine effectiveness of 35.8% (instead of 51.5%)* | Montréal | 14% (3%-31%) | 14% (12%-20%) | 14% (11%-22%) | 41% (29%-57%) | 28% (17%-46%) | 28% (16%-47%) | 27% (23%-40%) |
|  | Toronto | 11% (2%-26%) | 14% (12%-21%) | 14% (11%-28%) | 39% (28%-56%) | 24% (16%-39%) | 27% (16%-44%) | 26% (22%-46%) |
|  | Vancouver | 10% (2%-22%) | 14% (13%-17%) | 29% (25%-36%) | 49% (42%-59%) | 23% (16%-33%) | 39% (31%-49%) | 40% (35%-49%) |
| *Vaccine effectiveness of 86.0% (instead of 51.5%)* | Montréal | 19% (4%-43%) | 14% (11%-22%) | 38% (28%-55%) | 64% (50%-80%) | 34% (18%-55%) | 56% (41%-74%) | 48% (37%-66%) |
|  | Toronto | 14% (3%-25%) | 17% (12%-23%) | 56% (30%-66%) | 75% (52%-84%) | 29% (19%-40%) | 67% (43%-77%) | 66% (39%-77%) |
|  | Vancouver | 11% (3%-21%) | 12% (11%-15%) | 59% (52%-71%) | 74% (68%-83%) | 23% (17%-31%) | 69% (62%-79%) | 66% (59%-77%) |
| *Standardizing the start and coverage of vaccination* | Montréal | 15% (3%-34%) | 14% (12%-21%) | 41% (33%-57%) | 63% (52%-78%) | 29% (17%-48%) | 55% (42%-70%) | 51% (42%-68%) |
|  | Toronto | 11% (2%-27%) | 14% (12%-22%) | 38% (29%-60%) | 58% (46%-79%) | 25% (16%-40%) | 50% (37%-70%) | 47% (38%-72%) |
|  | Vancouver | 10% (2%-22%) | 14% (12%-16%) | 39% (35%-48%) | 58% (52%-69%) | 23% (16%-33%) | 50% (43%-60%) | 49% (44%-59%) |
| CrI: 95% Credible interval; VE: vaccine effectiveness.  ‡: model does not fit observed epidemic trajectory. | | | | | |  |  |  |

### Supplementary Figures

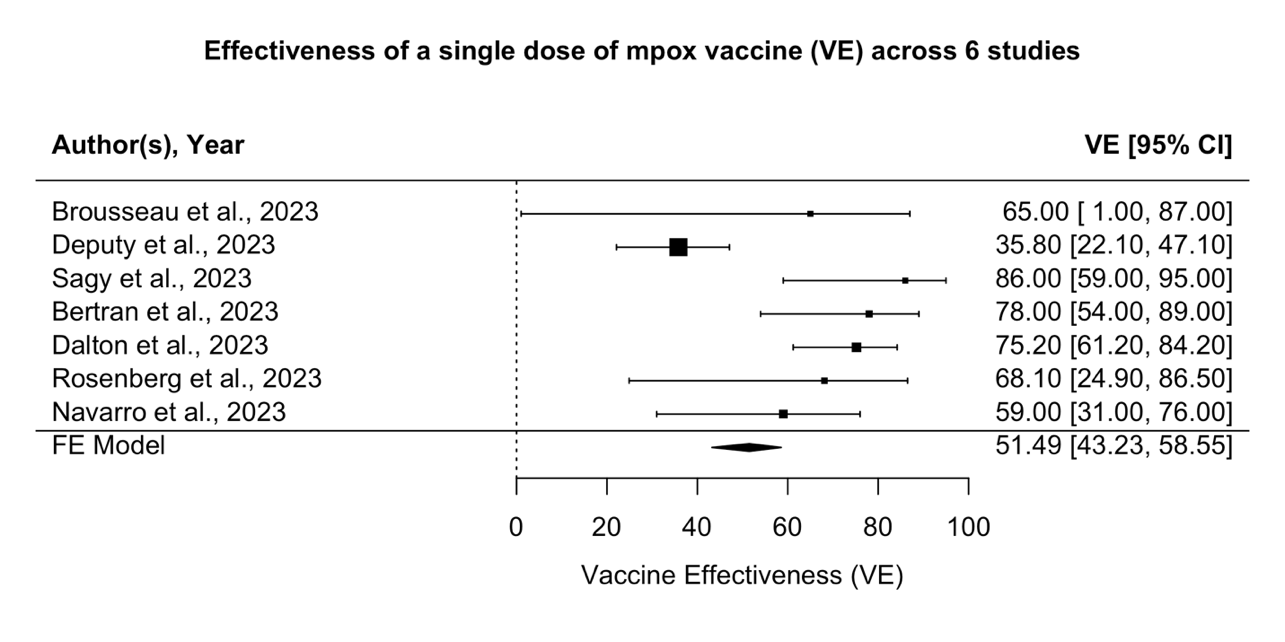

Figure S1. Effectiveness of a single dose of mpox vaccine (VE) across 7 studies. CI: confidence interval. Note that Sagy et al. estimated using hazard ratio, the rest of the studies used odds ratio. Squares are the mean and bars represent 95% confidence intervals.

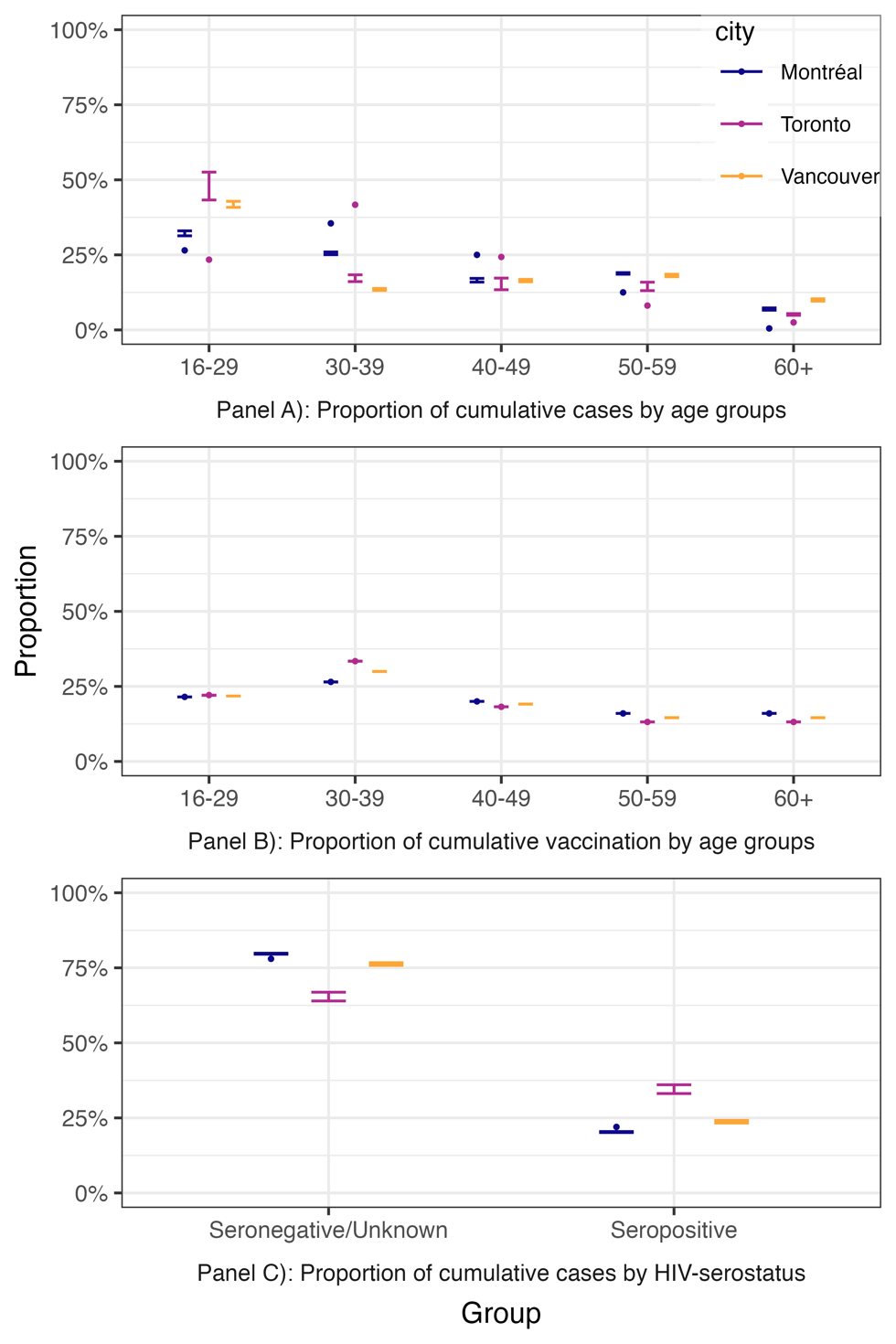

Figure S2. Cross-validation of model outcomes to available reported indicators**.** Modeled and observed proportion of cumulative cases A) age groups (not available from Vancouver), B) proportion of cumulative vaccinations by age groups (not available from Vancouver), and cumulative cases by HIV status (not available from Toronto and Vancouver) for each city. Bars represent the 95% credible intervals of model estimates, and points are the observed data. The lack of points for some cities and indicators indicate that the information is not available.

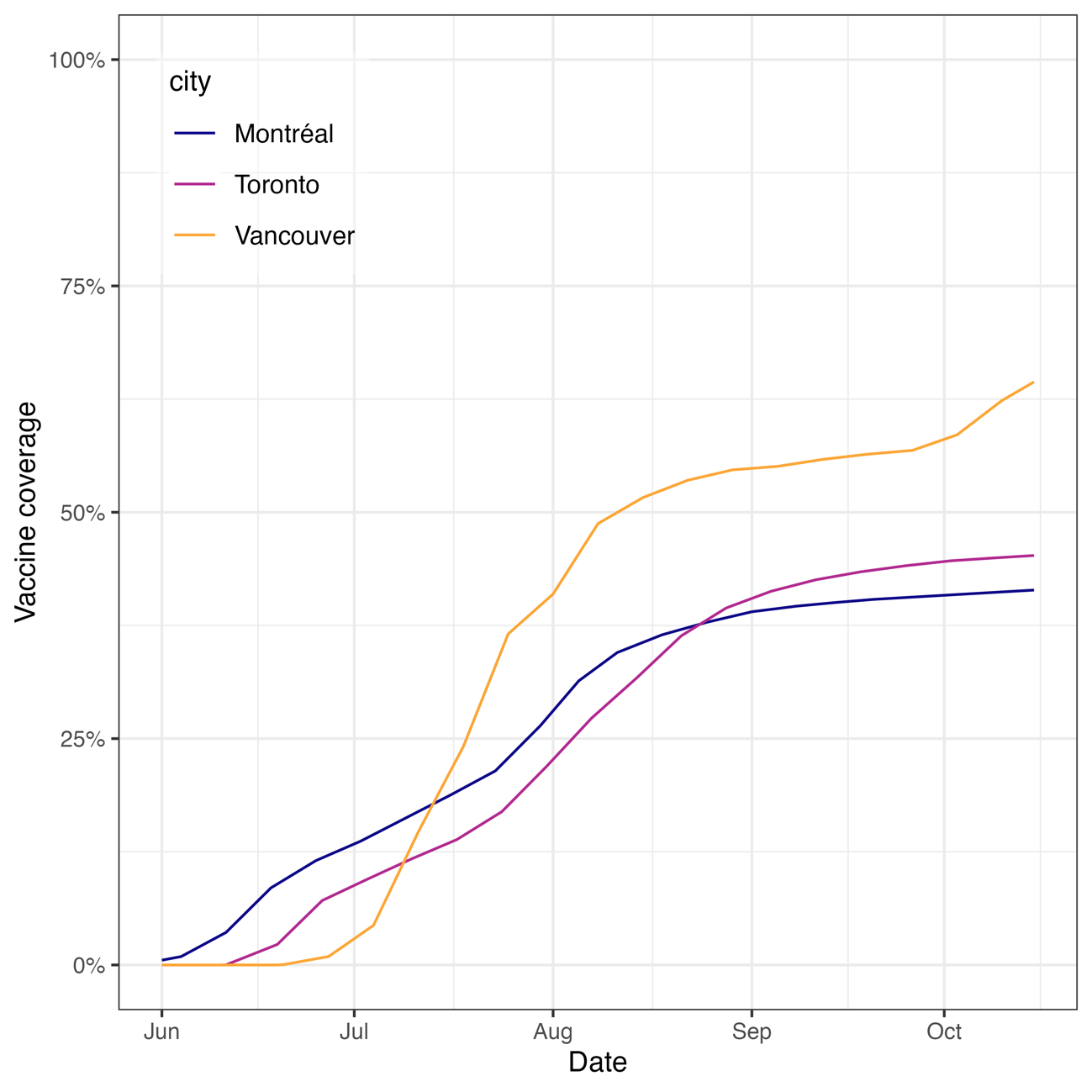

Figure S3. Estimated first-dose vaccine coverage of MVA-BN (Imvamune®) in Montréal, Toronto, and Vancouver from June 1^st^ to October 15^th^, 2022**.** Lines represent the estimated vaccine coverage across time using population sizes of gay, bisexual, and other men who have sex with men (denominators) and weekly-varying number of first doses (numerators). The city-specific number of first doses in Toronto was approximated by the provincial number of first doses in Ontario. For Vancouver, only the total number of first- and second-doses is available but few second-doses were administered before October 2022 in that city.
